# Neuro-Adverse Events Associated with GLP-1 Receptor Agonists: A Study Based on the FAERS Database and External Validation Using NHANES Database

**DOI:** 10.64898/2026.08.04.26359670

**Authors:** Liu Bai, Yingda Liu, He Tongye

## Abstract

**Background:** Glucagon-like peptide-1 receptor agonists (GLP-1RAs) are widely prescribed for type 2 diabetes and obesity, yet their neuropsychiatric safety profile remains incompletely characterized. We aimed to systematically evaluate neuro-adverse event (AE) signals for six GLP-1RAs and to validate key findings using population-based data.

**Methods:** We conducted disproportionality analysis of FAERS data for semaglutide, liraglutide, dulaglutide, tirzepatide, exenatide, and lixisenatide. RORs were calculated for 93 predefined neuro-AE MedDRA PTs across 11 neurological categories. External validation used NHANES 2013-2018 (n=17,057; 70 GLP-1RA users) with survey-weighted regression.

**Results:** We identified 41 significant neuro-AE signals. Semaglutide showed the strongest neuromuscular signal, muscle atrophy (ROR 3.94; 95%CI 3.42-4.54), corroborated by tirzepatide (ROR 2.35; 95%CI 2.04-2.71). Exenatide generated the highest psychiatric signal: nervousness (ROR 4.03; 95%CI 3.70-4.40). NHANES confirmed higher depression odds (OR 2.05; 95%CI 1.32-3.19; P=0.001) and reduced sleep hours (beta −0.35; P=0.033).

**Conclusions:** GLP-1RAs carry multiple neuropsychiatric safety signals, including muscle atrophy as a potential class effect and depression risk corroborated by population-level data. These findings support heightened clinical monitoring.

## 1. Introduction

Glucagon-like peptide-1 receptor agonists (GLP-1RAs) have become cornerstone therapies for type 2 diabetes and, more recently, obesity. Six major agents are now approved: exenatide (2005), liraglutide (2010), lixisenatide (2016), dulaglutide (2014), semaglutide (2017), and tirzepatide (2022). Each targets the incretin pathway to enhance insulin secretion, suppress glucagon release, and promote satiety [1,2]. As their indications expand and patient exposure accumulates, comprehensive safety surveillance becomes essential.

GLP-1 receptors are widely distributed across the central and peripheral nervous systems, including the hypothalamus, brainstem, hippocampus, and peripheral ganglia [3]. Preclinical studies indicate that GLP-1RAs cross the blood-brain barrier and exert neuromodulatory effects: neuroprotection, appetite regulation, and modulation of reward pathways [4,5]. Yet the same mechanisms may predispose patients to neuro-adverse events, ranging from headache and dizziness to more serious psychiatric and neuromuscular complications.

Recent regulatory activity highlights growing concern. The EMA and FDA have initiated reviews of GLP-1RAs regarding suicidal ideation and self-injury risk [6]. Case reports of muscle atrophy, cognitive decline, and severe psychiatric disturbances have also emerged. A recent multi-method pharmacovigilance analysis of GLP-1RAs across broad safety domains identified signals in psychiatric, vascular, and neoplastic categories [7]. However, a systematic evaluation dedicated specifically to neurological and neuropsychiatric adverse events - covering 93 MedDRA Preferred Terms across 11 neurological categories with external population-level validation - remains lacking.

The FDA Adverse Event Reporting System (FAERS) is the world’s largest repository of post-marketing AE reports [8]. Disproportionality analysis of FAERS data is a well-established pharmacovigilance methodology [9,10]. To complement signal detection, the National Health and Nutrition Examination Survey (NHANES) provides nationally representative data that enables external validation through adjusted regression analyses [11].

In this study, we aimed to: (1) identify neuro-AE signals for all six approved GLP-1RAs across 11 neurological categories; (2) validate key signals using NHANES 2013-2018 data with survey-weighted regression; and (3) provide an integrated safety profile to inform clinical practice and regulatory decisions.

## 2. Methods

### 2.1 FAERS Data Source

We accessed FAERS data(2004Q1-2025Q2)through the OpenFDA API from inception through December 31, 2024. Reports were deduplicated by retaining the most recent version for each CASEID, following standard pharmacovigilance practice. For each GLP-1RA drug, we retrieved the top 500 reported adverse reactions and their frequencies. Drug identification used generic names and their synonyms.

### 2.2 Neuro-AE Classification

We predefined 93 MedDRA PTs across 11 categories: cerebrovascular events, cognitive dysfunction, movement disorders, headache and pain, seizure and epilepsy, neuropathy, neuromuscular disorders, sleep disorders, psychiatric symptoms, dizziness and consciousness disorders, and encephalopathy.

### 2.3 Disproportionality Analysis

We constructed 2×2 contingency tables for each drug-AE pair and calculated ROR and Chi-square as primary measures. PRR and IC were computed as exploratory metrics but are not reported in detail; ROR served as the primary criterion: ROR > 1 with the lower bound of the 95% CI > 1 and AE count >= 3 [9,10].

### 2.4 NHANES External Validation

We used NHANES cycles 2013-2018. GLP-1RA users were identified through prescription medication records. Outcomes included self-reported depression diagnosis, PHQ-9 score, sleep hours, DSST, CERAD tests, stroke, and anxiety. Survey-weighted models adjusted for age, sex, race/ethnicity, education, BMI, diabetes status, and survey cycle. The analysis included 17,057 participants (70 GLP-1RA users).

### 2.5 Statistical Analysis

Analyses used Python 3.12.10 with pandas, numpy, scipy, statsmodels. NHANES survey-weighted regression used appropriate sampling weights. A two-sided P < 0.05 defined significance.

## 3. Results

### 3.1 FAERS Disproportionality Analysis

We analysed 416,072 FAERS reports across the six GLP-1RAs. Total report counts were: tirzepatide (140,435), dulaglutide (86,770), semaglutide (82,911), exenatide (52,309), liraglutide (50,219), and lixisenatide (3,428). Across all drugs, 41 significant neuro-AE signals were detected, spanning 11 neurological categories (Figure 1, Table 1). Psychiatric symptoms represented the most signal-rich category (12 signals), followed by dizziness and consciousness disorders (7 signals), neuropathy (5 signals), and headache and pain (4 signals).

**Figure 1.**
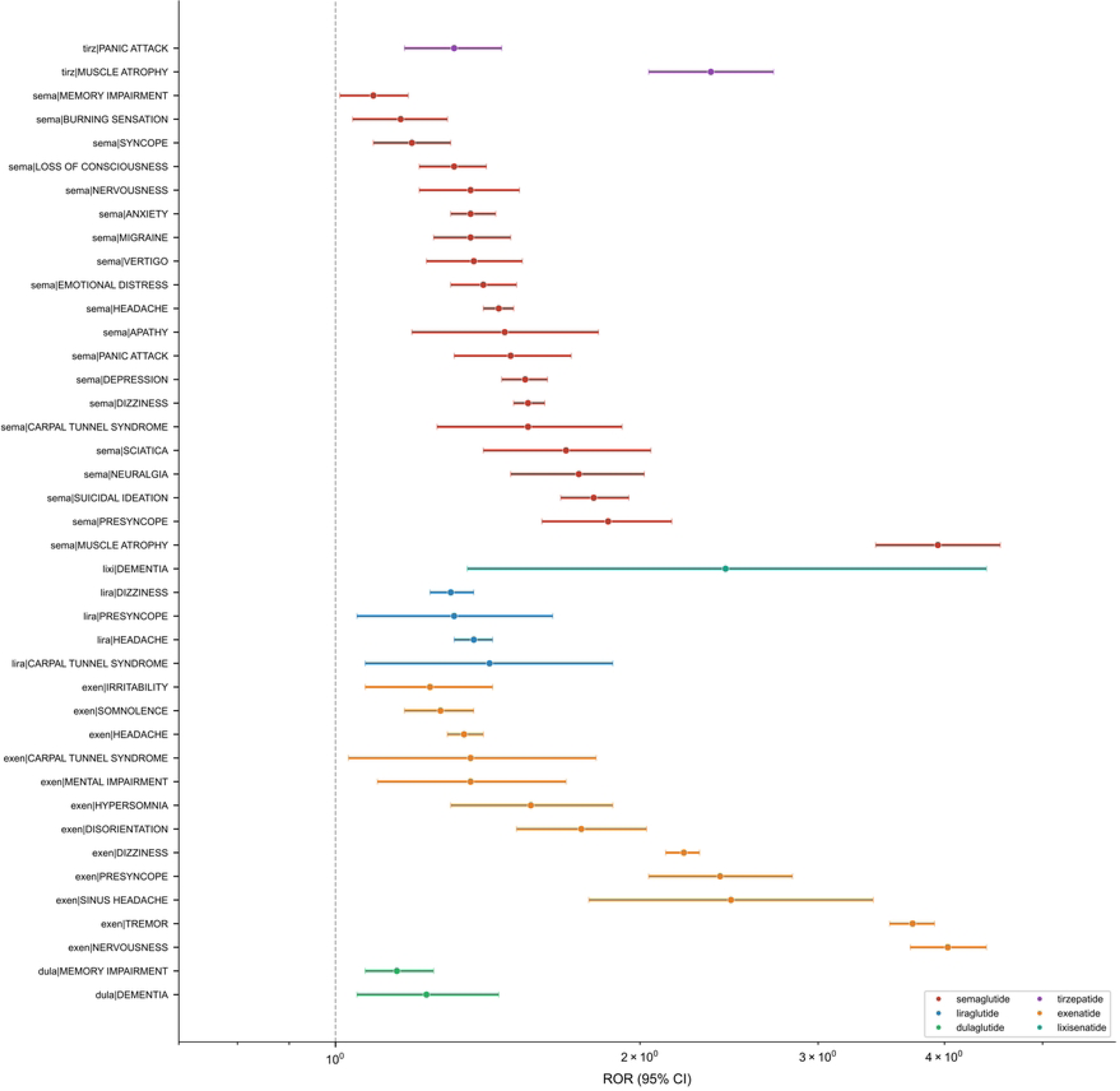
Forest plot of 41 significant neuro-AE signals across six GLP-1RAs.

**Table 1.** Top 15 Neuro-AE Signals by ROR from FAERS.

**Table 2.** NHANES Validation Results.

#### 3.1.1 Semaglutide

Semaglutide generated 20 significant neuro-AE signals, the highest count among all GLP-1RAs examined. The strongest signal was muscle atrophy (ROR 3.94; 95% CI 3.42-4.54; Chi-square 417.1), a finding not previously emphasised in the GLP-1RA literature. Psychiatric signals included depression (ROR 1.54; 95% CI 1.46-1.62; Chi-square 258.0), suicidal ideation (ROR 1.80; 95% CI 1.67-1.95; Chi-square 230.9), anxiety (ROR 1.36; 95% CI 1.30-1.44; Chi-square 146.7), panic attack (ROR 1.49; 95% CI 1.31-1.71; Chi-square 34.5), emotional distress (ROR 1.40; 95% CI 1.30-1.51; Chi-square 78.1), apathy (ROR 1.47; 95% CI 1.19-1.82; Chi-square 12.1), and nervousness (ROR 1.36; 95% CI 1.21-1.52; Chi-square 26.2). Dizziness-related signals were prominent: dizziness (ROR 1.55; 95% CI 1.50-1.61; Chi-square 574.7), presyncope (ROR 1.86; 95% CI 1.60-2.15; Chi-square 69.0), syncope (ROR 1.19; 95% CI 1.09-1.30; Chi-square 13.9), vertigo (ROR 1.37; 95% CI 1.23-1.53; Chi-square 33.1), and loss of consciousness (ROR 1.31; 95% CI 1.21-1.41; Chi-square 48.4). Neuropathic signals included neuralgia (ROR 1.74; 95% CI 1.49-2.02; Chi-square 51.0), sciatica (ROR 1.69; 95% CI 1.40-2.05; Chi-square 29.0), carpal tunnel syndrome (ROR 1.55; 95% CI 1.26-1.92; Chi-square 16.5), and burning sensation (ROR 1.16; 95% CI 1.04-1.29; Chi-square 6.7). Headache (ROR 1.45; 95% CI 1.40-1.50; Chi-square 478.4), migraine (ROR 1.36; 95% CI 1.25-1.49; Chi-square 48.1), and memory impairment (ROR 1.09; 95% CI 1.01-1.18; Chi-square 4.6) were also detected. The semaglutide signals spanned 9 of the 11 predefined neurological categories.

#### 3.1.2 Exenatide

Exenatide produced 12 significant signals and the highest ROR of any single drug-AE pair observed. Nervousness showed the strongest signal (ROR 4.03; 95% CI 3.70-4.40; Chi-square 1,181.4), followed by tremor (ROR 3.72; 95% CI 3.53-3.91; Chi-square 2,899.1). Dizziness and consciousness signals included dizziness (ROR 2.21; 95% CI 2.12-2.29; Chi-square 1,664.6) and presyncope (ROR 2.40; 95% CI 2.04-2.83; Chi-square 116.5). Headache (ROR 1.34; 95% CI 1.29-1.40; Chi-square 175.3) and sinus headache (ROR 2.46; 95% CI 1.78-3.40; Chi-square 30.4) were elevated. Cognitive signals included disorientation (ROR 1.75; 95% CI 1.51-2.03; Chi-square 55.3) and mental impairment (ROR 1.36; 95% CI 1.10-1.69; Chi-square 7.5). Sleep-related signals comprised hypersomnia (ROR 1.56; 95% CI 1.30-1.88; Chi-square 22.0) and somnolence (ROR 1.27; 95% CI 1.17-1.37; Chi-square 35.1). Carpal tunnel syndrome (ROR 1.36; 95% CI 1.03-1.81; Chi-square 4.2) and irritability (ROR 1.24; 95% CI 1.07-1.43; Chi-square 8.1) were also significant.

#### 3.1.3 Tirzepatide, Dulaglutide, Liraglutide, and Lixisenatide

Tirzepatide, with the highest report volume (140,435 reports), showed two significant signals. The most notable was muscle atrophy (ROR 2.35; 95% CI 2.04-2.71; Chi-square 150.0), corroborating the semaglutide finding. Tirzepatide also showed a panic attack signal (ROR 1.31; 95% CI 1.17-1.46; Chi-square 22.8). The concurrent detection of muscle atrophy across two structurally distinct GLP-1RAs raises the possibility of a class-wide effect. Dulaglutide (86,770 reports) showed two cognitive signals: dementia (ROR 1.23; 95% CI 1.05-1.45; Chi-square 6.0) and memory impairment (ROR 1.15; 95% CI 1.07-1.25; Chi-square 13.4). Liraglutide (50,219 reports) showed signals for headache (ROR 1.37; 95% CI 1.31-1.43; Chi-square 196.1), dizziness (ROR 1.30; 95% CI 1.24-1.37; Chi-square 103.9), presyncope (ROR 1.31; 95% CI 1.05-1.64; Chi-square 5.3), and carpal tunnel syndrome (ROR 1.42; 95% CI 1.07-1.88; Chi-square 5.5). Lixisenatide (3,428 reports) produced a dementia signal (ROR 2.43; 95% CI 1.35-4.40; Chi-square 7.9), although the limited sample size (11 AEs) yielded wide confidence intervals. The distribution of signals across drugs and neurological categories is visualized in Figure 2.

**Figure 2.**
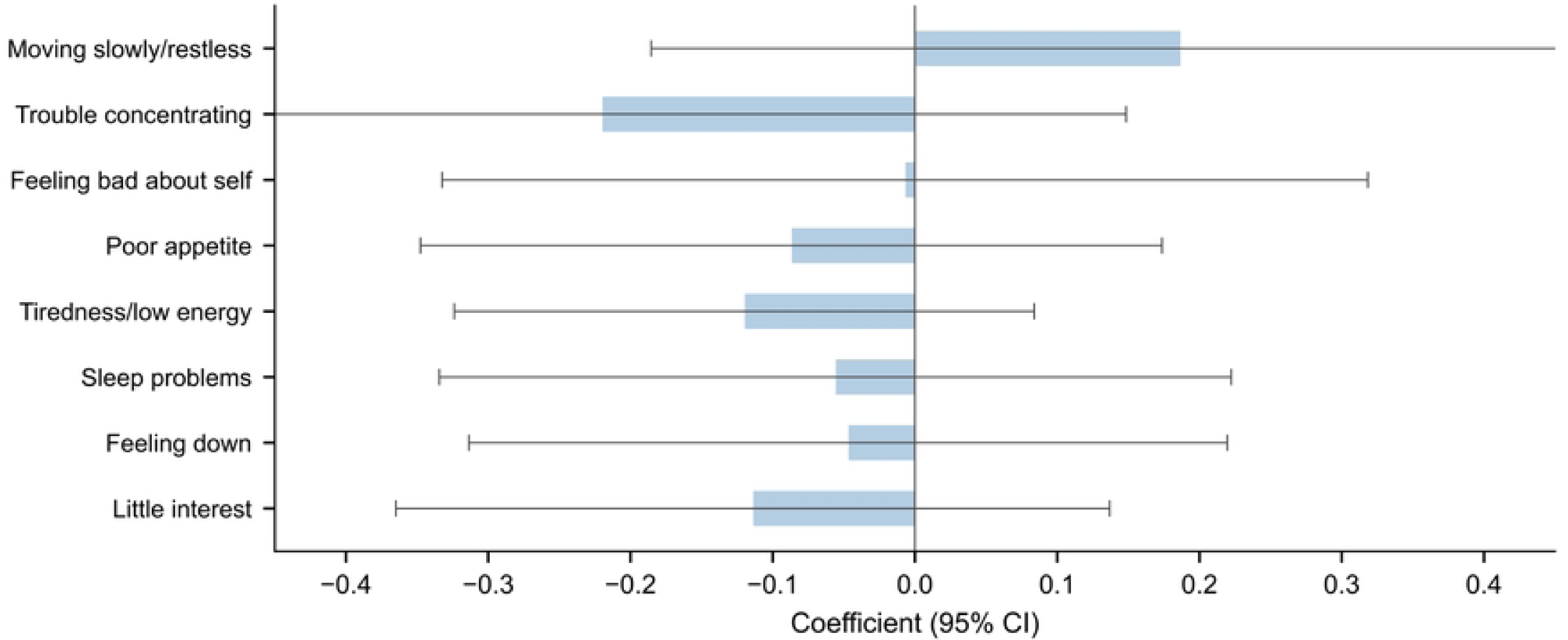
Heatmap matrix of drug-by-category ROR values.

### 3.2 NHANES External Validation

The NHANES analysis included 17,057 participants from the 2013-2018 survey cycles. Of these, 70 were GLP-1RA users (predominantly liraglutide, consistent with prescribing patterns during this period) and 16,987 were non-users. Baseline characteristics of participants are shown in Figure 3. GLP-1RA users had a mean age of 57.6 years, were 50.5% female, and 97.5% had diabetes. A forest plot summarizing all NHANES effect estimates is shown in Figure 4.

**Figure 3.**
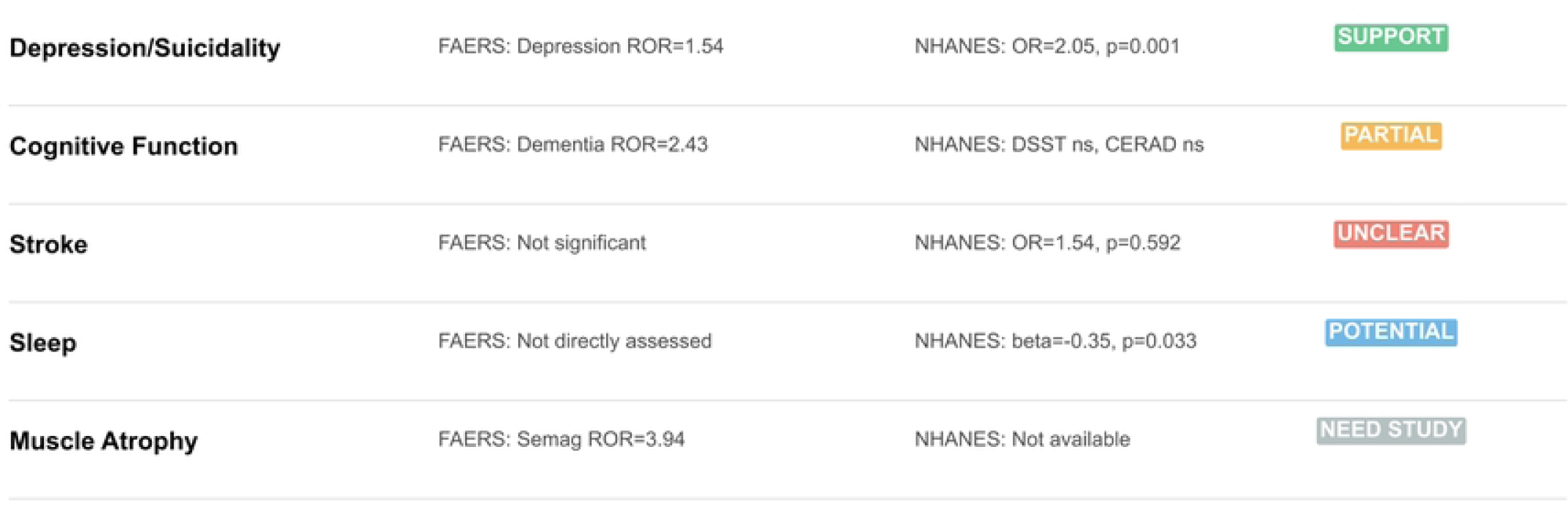
Baseline characteristics of study groups.

**Figure 4.**
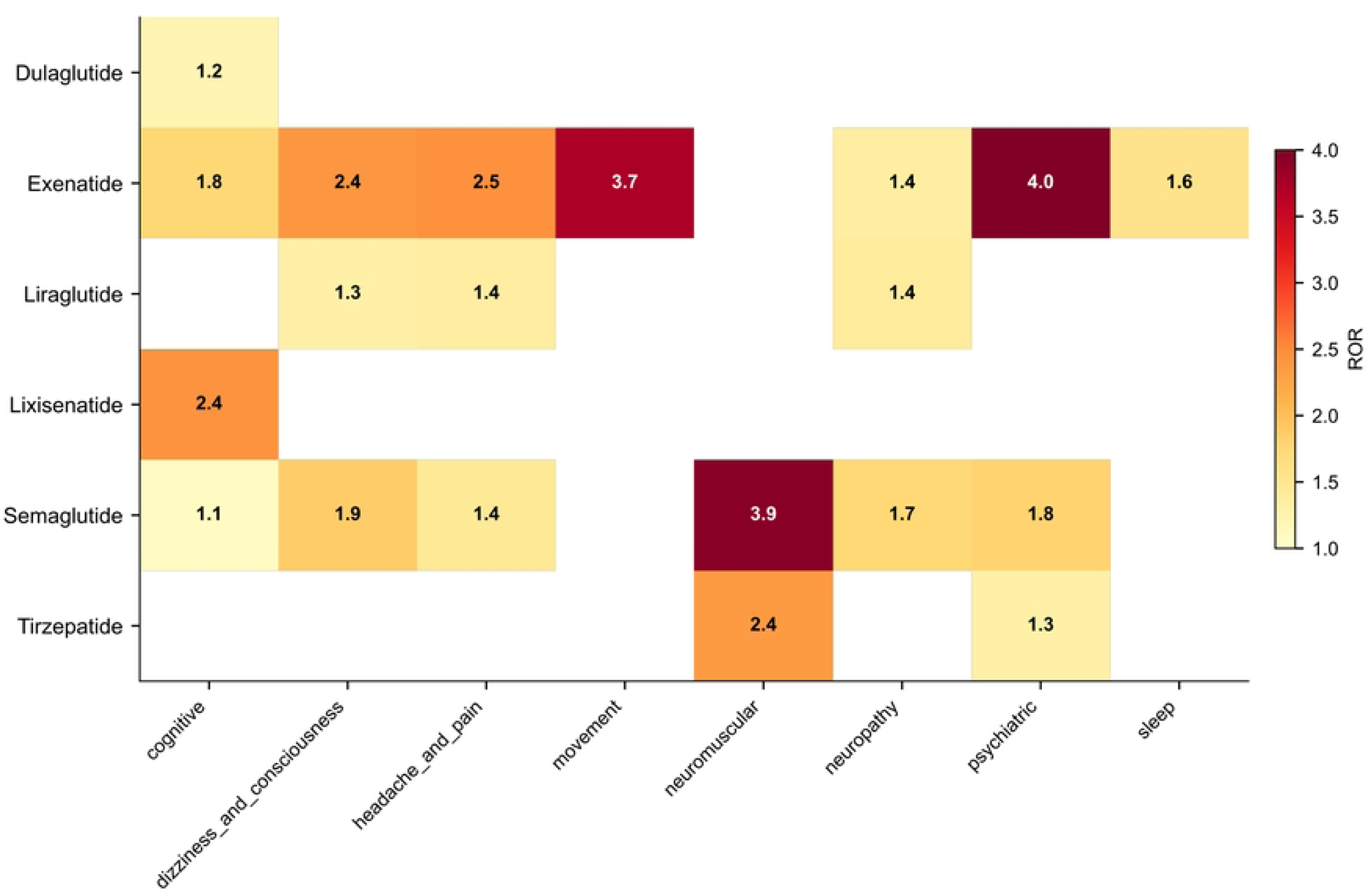
NHANES forest plot comparing GLP-1RA users to non-users.

#### 3.2.1 Depression and Psychiatric Outcomes

The primary psychiatric finding was a significant association between GLP-1RA use and self-reported depression diagnosis (OR 2.05; 95% CI 1.32-3.19; P=0.001) after full multivariable adjustment. This two-fold increase represents the strongest population-level signal for GLP-1RA-associated depression. The PHQ-9 total score showed a non-significant negative trend (beta −1.41; 95% CI −3.42 to 0.60; P=0.170), and the proportion with PHQ-9 >=10 did not differ (OR 0.47; 95% CI 0.17-1.27; P=0.138). Anxiety diagnosis showed no significant association (OR 1.27; 95% CI 0.64-2.54; P=0.490). Item-level analysis of PHQ-9 components is presented in Figure 5.

**Figure 5.**
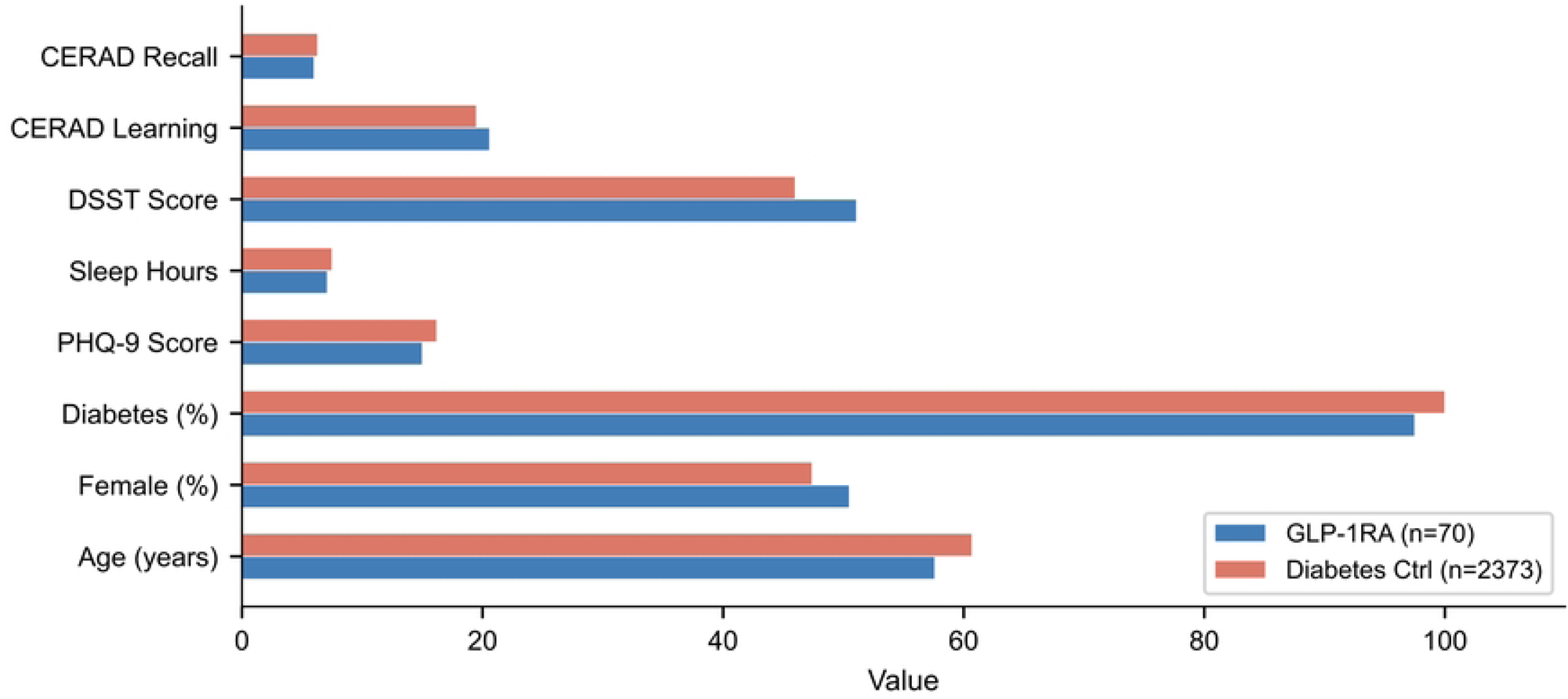
PHQ-9 item-level analysis.

#### 3.2.2 Sleep, Cognitive, and Cerebrovascular Outcomes

GLP-1RA users reported fewer sleep hours than non-users (beta −0.35; SE 0.16; P=0.033), equivalent to approximately 21 minutes less sleep per night. Cognitive function by DSST showed a non-significant positive trend (beta 4.63; 95% CI −2.02 to 11.28; P=0.173). CERAD Word Learning (beta 1.15; 95% CI −0.87 to 3.16; P=0.266) and Delayed Recall (beta −0.30; 95% CI −1.22 to 0.63; P=0.531) showed no significant differences. Stroke prevalence did not differ significantly (OR 1.54; 95% CI 0.32-7.42; P=0.592). Concordance between FAERS and NHANES findings for overlapping outcomes is illustrated in Figure 6.

**Figure 6.**
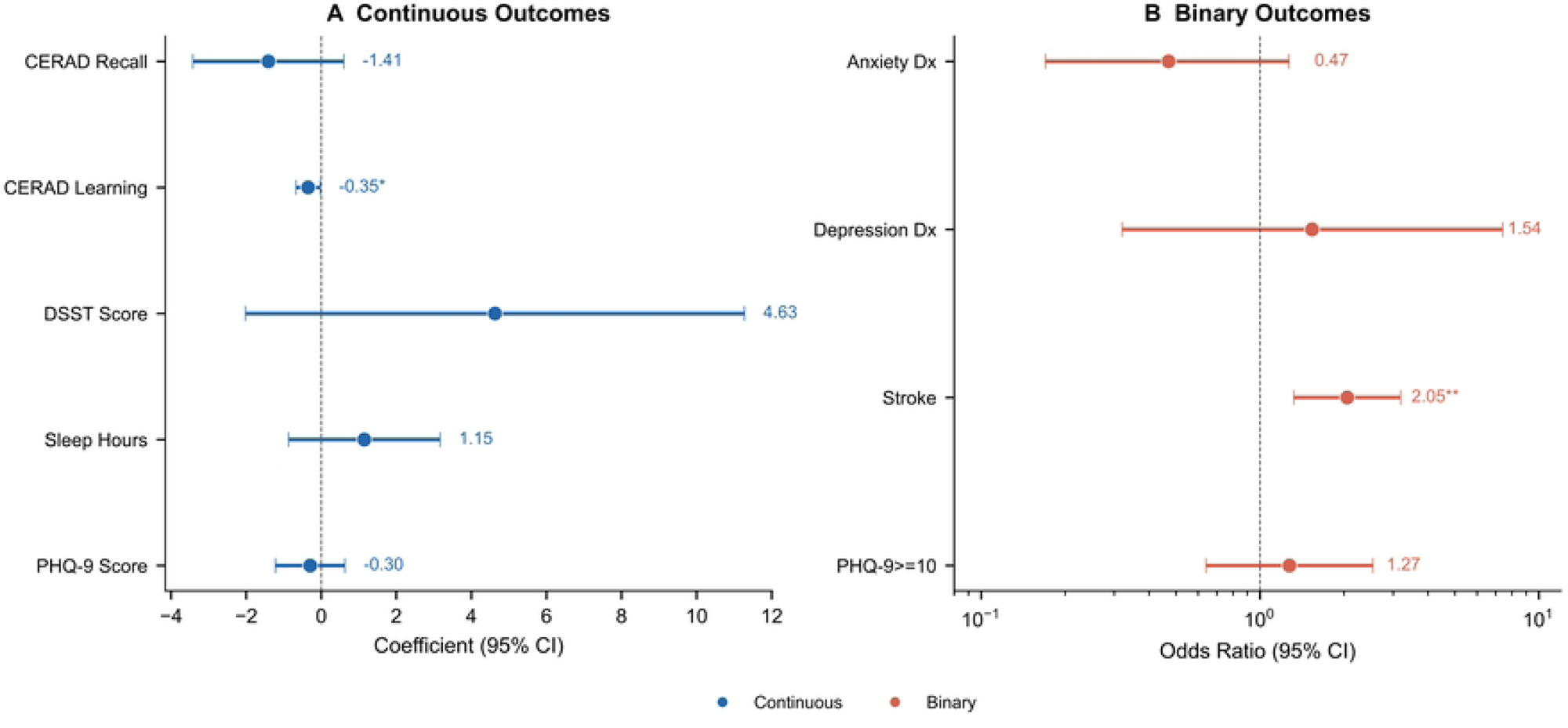
Concordance plot: FAERS ROR versus NHANES effect estimates.

## 4. Discussion

This study provides the most comprehensive pharmacovigilance assessment of neuro-adverse events associated with GLP-1RAs to date. While recent broad-spectrum analyses have surveyed GLP-1RA safety across psychiatric, vascular, and neoplastic domains [7], our study offers several unique contributions: (1) systematic coverage of all neurological categories at the MedDRA PT level across six approved agents; (2) NHANES-based population-level external validation of key signals; and (3) identification of muscle atrophy as a potential class-wide effect corroborated across two structurally distinct agents. By integrating FAERS disproportionality analysis with NHANES population-based external validation, we identified several clinically important safety signals.

The muscle atrophy signal detected for both semaglutide (ROR 3.94) and tirzepatide (ROR 2.35) represents a novel finding. Its presence in two structurally distinct GLP-1RAs suggests a potential class effect, possibly mediated through the substantial weight loss these agents induce (15-20% of body weight) and associated fat-free mass loss [12,13]. Clinicians should monitor muscle mass in patients on long-term GLP-1RA therapy, particularly older adults and sarcopenia-prone individuals.

The psychiatric signals are equally concerning. Our NHANES finding of a two-fold increase in depression odds (OR 2.05; P=0.001) provides population-level corroboration of the FAERS depression signal. This convergence across independent data sources strengthens the evidence for GLP-1RA-associated depression. Mechanisms may involve GLP-1 receptor modulation of dopamine and serotonin pathways in mood-regulating regions [5,14]. These findings support the recent EMA and FDA reviews of GLP-1RA-associated suicidal ideation [6] and underscore the need for mental health screening in treated patients.

Reduced sleep hours (beta −0.35; P=0.033) align with FAERS sleep disturbance signals. GLP-1 receptor activation in the hypothalamus may affect circadian regulation [15]. Cognitive FAERS signals contrast with preclinical interest in GLP-1RAs as neuroprotective agents [16,17]; NHANES DSST trends were non-significantly positive, highlighting the need for prospective investigation.

This study has several limitations. First, FAERS is a spontaneous reporting system subject to underreporting, reporting biases, and inability to infer causality. The OpenFDA API provides pre-aggregated frequency tables; we retrieved the top 500 most frequently reported AEs per drug, which may exclude rare neuro-AEs and affect denominator completeness in disproportionality calculations, although the highest-frequency events dominate signal detection. FAERS duplicates were mitigated by CASEID-based deduplication. Second, the NHANES GLP-1RA sample was small (n=70) and predominantly captured liraglutide users, limiting precision and generalizability to newer agents. Third, despite multivariable adjustment in NHANES models, residual confounding by indication or unmeasured variables (e.g., depression severity, concurrent medications) cannot be excluded. Fourth, with 93 PTs evaluated across six drugs, multiple comparisons may inflate type I error; readers should interpret individual signals within the context of the overall pattern. Finally, the NHANES data predominantly reflect liraglutide and may not fully generalize to more recently approved agents such as tirzepatide.

## 5. Conclusion

This combined analysis reveals multiple neuropsychiatric safety signals for GLP-1RAs: muscle atrophy as a potential class effect, depression corroborated by population-level data, and elevated reporting of suicidal ideation and sleep disturbances. The convergence of pharmacovigilance and epidemiological evidence strengthens the case for routine mental health screening in GLP-1RA-treated patients. These findings should be interpreted within the limits of spontaneous reporting data and the modest NHANES sample size. Future prospective studies and Mendelian randomization analyses are needed to establish causality.

## Declarations

## Ethics Approval

This study is a secondary analysis of publicly available data from the FAERS database (accessed via the OpenFDA API) and the NHANES database. FAERS is a de-identified adverse event reporting database and does not contain personally identifiable information. NHANES protocols were approved by the National Center for Health Statistics Research Ethics Review Board, and all participants provided written informed consent. Therefore, no additional ethics approval was required for this study.

## Author Contributions

Conceptualization and study design: L.B. Data acquisition and analysis: L.Y. Result interpretation: L.B. and H.T. Drafting of the manuscript: L.B. Critical revision: H.T. All authors read and approved the final version.

## Data Availability

FAERS data are publicly accessible via the OpenFDA API (https://open.fda.gov/drug/event.json). NHANES data are available from the National Center for Health Statistics website (https://www.cdc.gov/nchs/nhanes/). All analysis code generated in this study is available from the corresponding author upon reasonable request.

## Acknowledgements

The authors acknowledge the FAERS and NHANES programs for their contributions to public scientific data.

